# Undocumented infectives in the Covid-19 pandemic

**DOI:** 10.1101/2020.07.09.20149682

**Authors:** Maurizio Melis, Roberto Littera

**Author notes:** The authors contributed equally to this work.

## Abstract

**Background:** A crucial role in epidemics is played by the number of undetected infective individuals who continue to circulate and spread the disease. Epidemiological investigations and mathematical models have revealed that the rapid diffusion of Covid-19 can mostly be attributed to the large percentage of undocumented infective individuals who escape testing.

**Methods:** The dynamics of an infection can be described by the SIR model, which divides the population into susceptible (*S*), infective (*I*) and removed (*R*) subjects. In particular, we exploited the Kermack and McKendrick epidemic model which can be applied when the population is much larger than the fraction of infected subjects.

**Results:** We proved that the fraction of undocumented infectives, in comparison to the total number of infected subjects, is given by 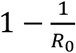, where *R*_0_ is the basic reproduction number. The mean value *R*_0_ = 2.10 (2.09 − 2.11) for the Covid-19 epidemic in three Italian regions yielded a percentage of undetected infectives of 52.4% (52.2% - 52.6%) compared to the total number of infectives.

**Conclusions:** Our results, straightforwardly obtained from the SIR model, highlight the role played by undetected carriers in the transmission and spread of the SARS-CoV-2 infection. Such evidence strongly recommends careful monitoring of the infective population and ongoing adjustment of preventive measures for disease control until a vaccine becomes available.

## Introduction

A critical issue in the control of an epidemic is to know the exact number of infective subjects. Current estimates of SARS-CoV-2 infection are significantly hampered by the difficulty to perform large-scale diagnostic tests, despite a growing awareness that the spread of the Covid-19 pandemic is mostly caused by undetected carriers.

The dynamics of an epidemic can be described by an epidemiological model known as the SIR model, which divides the whole population into three classes of subjects: susceptible (*S*), infective (*I*) and removed (*R*) individuals. Kermack and McKendrick [1] developed a SIR model for the study of epidemics in populations much larger than the infected fraction. Under this assumption, which is fully verified in the Covid-19 epidemic, we proved that the total number of infectives, when an epidemic occurs, is approximately *R*_0_·*R*, where *R*_0_ > 1 is the basic reproduction number of the infection and *R* is the number of infectives who have been removed because of recovery, isolation, hospitalisation or death. The number of undocumented infectives is then (*R*_0_ − 1)·*R*. The fractions of removed and undetected infectives, in comparison to the total number of infectives, are 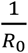 and 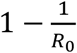, respectively.

By applying the aforesaid model to the data available on the Covid-19 epidemic in Italy, we obtained that the mean value of the basic reproduction number in three Italian regions was *R*_0_ = 2.10 (95% confidence interval, 2.09 – 2.11). Consequently, the number of undocumented cases turned out to be about *R*_0_ − 1 = 1.1 times the number of removed cases. More specifically, we found that the percentage of undocumented infectives was about 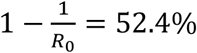 (95% confidence interval, 52.2% – 52.6%) of the total number of infectives.

Previous investigations found that the percentages of asymptomatic infectives (i.e. subjects without fever, cough or any other symptoms) were: 43.2% (32.2% - 54.7%) in Vo’, a small town near Padua in Italy [2]; 50.5% (46.5% - 54.4%) on board the Diamond Princess cruise ship in Yokohama, Japan [3]; 47% (38% - 56%) in mainland China [4] and 52.0% (including paucisymptomatic infectives) in a large sample (64660 subjects) of the Italian population [5].

The speed at which an epidemic grows cannot be explained if we only take into account the number of recorded infected patients who, supposably, are immediately removed from the circulating population by hospitalisation or isolation at home.

Undocumented infectives are largely responsible for the rapid increase of the epidemic and can be classified into three classes: 1) paucisymptomatic or asymptomatic individuals, who never develop overt symptoms during the course of infection; 2) presymptomatic subjects, who will eventually develop symptoms; 3) symptomatic infective individuals, who have clinical symptoms but for several reasons (such as the shortage of nasopharyngeal swabs) are not diagnosed as positive.

The third category of infectives, if quarantined, do not transmit the disease and can only be detected by subsequent serological investigation. Our model can only reveal the subjects who actually contribute to the spread of the coronavirus disease, hence excluding the third class of infectives.

The undocumented infectives in the first and second categories (paucisymptomatic or asymptomatic carriers and presymptomatic individuals) continue to circulate and transmit the disease. To reliably detect their presence, it would be necessary to test the entire population and not just the symptomatic cases.

The data provided by the Italian Ministry of Health and the Civil Protection Department up to the 3^rd^ of June 2020 [6] reported about 233800 removed cases in Italy, including either patients hospitalised or isolated at home or recovered or dead. Based on the result found in the present study, the total number of paucisymptomatic, asymptomatic and presymptomatic infectives had to be almost 491000 up to that date. This means that 257200 individuals were not diagnosed as infected although they continued to circulate and spread the virus.

This study confirms that undocumented infectives can be considered the key culprits for the rapid spread of SARS-CoV-2 within the population. Consequently, interventions to control the infection will need to be maintained until the complete disappearance of the epidemic.

Further details on the SIR model and the numerical fit of the data are reported in the Appendices, where evaluation of the basic and effective reproduction numbers *R*_0_ and *R*_eff_(*t*) is also discussed.

## Methods

In the SIR epidemic model the population is divided into three distinct classes [7]: the susceptible subjects, *S*, who can catch the disease; the unremoved infectives, *I*, who have the disease and can transmit it; and the removed infected subjects, *R*, namely those with a laboratory diagnosis who are either hospitalised, isolated at home, dead or recovered.

We assume that all the individuals diagnosed as infected – either by nasopharyngeal swab or serological test – are immediately isolated, thus passing from the class of infectives *I* to that of the removed infectives *R*. On the contrary, the infected subjects with a positive diagnosis are classified as undocumented infectives (*U*), who are either still infective (*I*) or infected but no longer contagious (*U*_0_). The total number of undocumented infectives *U* is then given by: *U* = *I* + *U*_0_.

At any time *t*, the total number of infected subjects *I_tot_*(*t*) is the sum of the number of removed infectives *R*(*t*) and undocumented infectives *U*(*t*): *I_tot_*(*t*) = *R*(*t*)+*U*(*t*). As discussed in the Introduction, the undocumented infective individuals revealed by our model can be asymptomatic, paucisymptomatic or presymptomatic.

The progression of an individual from the susceptible compartment *S* to the total infected class *I_tot_* is represented by the scheme in Figure 1.

**Figure 1.**
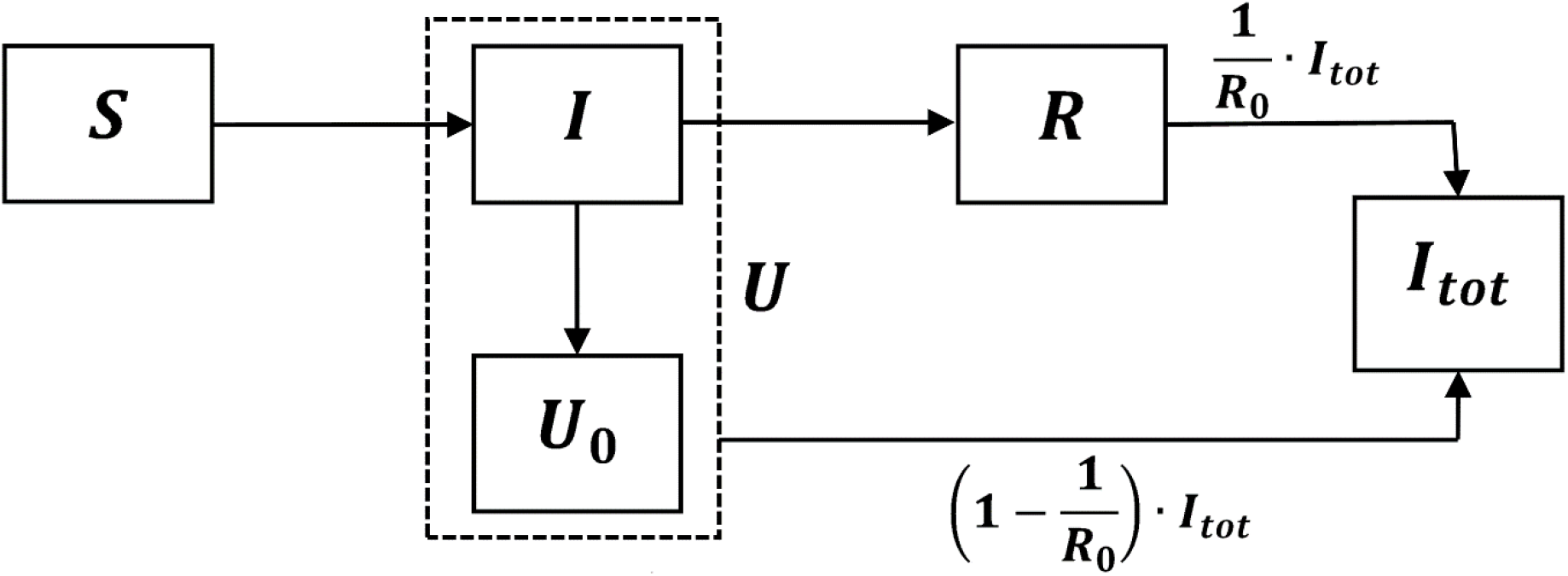
Scheme representing the progression of an individual from the susceptible compartment *S* to the total infected class *I_tot_*. The fractions of *I_tot_* in the *R* and *U* compartments are 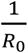 and 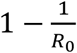, respectively.

If *S*(*t*) is the number of susceptible individuals at time *t* and *N* is the size of the population, the total number of infected subjects *I_tot_*(*t*) turns out to be

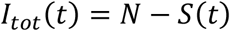

By manipulating the differential equations which define the SIR model (Appendix A) and assuming that the initial number *S*_0_ of susceptible individuals is close to *N*, i.e. *S*_0_ ≅ *N*, one obtains

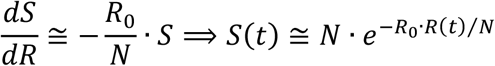

where *R*_0_ is the basic reproduction number (discussed in Appendix B). Under the assumption *R*_0_·*R*(*t*)*N/* ≪ 1 (a condition which is certainly verified if the population size is much larger than the number of infected subjects) we can approximate *S*(*t*) in the following form:

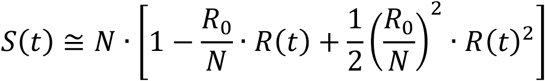

The total number of infected subjects *I_tot_*(*t*) at time *t* then becomes

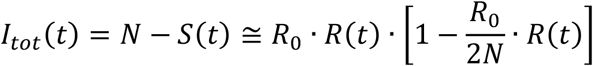

while the unremoved infectives *U*(*t*) at time *t* turn out to be

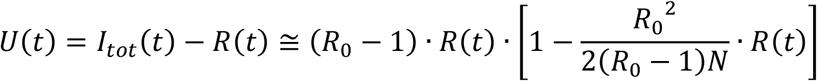

The ratio between the removed infected subjects *R*(*t*) and *I_tot_*(*t*) at time *t* is

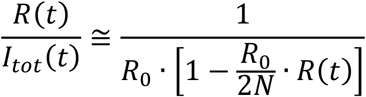

while the ratio between the unremoved infectives *U*(*t*) and *I_tot_*(*t*) at time *t* is

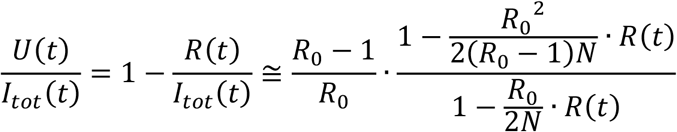

Being *R*_0_·*R*(*t*)*N/* ≪ 1, the previous four equations can be approximated as

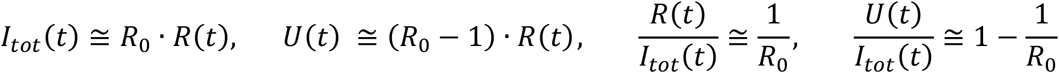

These results are obtained under the assumption *S*_0_ ≅ *N*, which implies *R*_0_ > 1, i.e. that an epidemic ensues.

The fraction of undocumented infectives *U*, in comparison to the total infectives *I_tot_*, has been derived straightforwardly from the SIR epidemic model and only depends on the basic reproduction number *R*_0_.

## Results

The data provided by the Italian Ministry of Health and the Civil Protection Department in Italy [6], updated to the 3^rd^ of June 2020, were fitted for three Italian regions by means of a specific code written with Wolfram Mathematica 12.1 [8] and based on the Kermack-McKendrick model [1].

Lombardy, in the north of Italy, has been the region with the highest number of Covid-19 infections, followed by Emilia-Romagna (at the second place from the 29^th^ of February to the 24^th^ of April, at the third place in the other periods of the epidemic). On the contrary, the Island of Sardinia, in the South of Italy, was one of the regions with the lowest number of documented Covid-19 infections and deaths. The population size *N* in these regions, updated to the 1st of January 2019, were: Lombardy *N* = 10060574, Emilia-Romagna *N* = 4459477, Sardinia *N* = 1639591 (data from ISTAT, Italian National Institute of Statistics).

In these three Italian regions our epidemiological model yielded the mean value *R*_0_ = 2.10 (2.09 − 2.11) for the basic reproduction number *R*_0_ (Appendix B).

Time *t* was expressed in days since *t*_0_ (*t* = 0), the day before the date of the first diagnosed patient: 19^th^ of February in Lombardy, 20^th^ of February in Emilia-Romagna and 2nd of March in Sardinia.

At any time *t*, the mean percentage of removed infectives *R*(*t*) in comparison to the total number of infectives *I_tot_*(*t*) was about 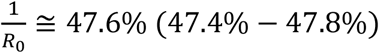, while the mean percentage of unremoved infectives *U*(*t*) was about 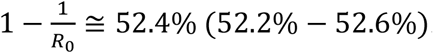.

Figure 2 represents, on the basis of the data provided by the Italian Ministry of Health [6], the number of removed infectives *R*(*t*) in Lombardy, Emilia-Romagna and Sardinia, fitted by the equation *R*(*t*) = *c*_1_·[tanh(*c*_2_*t* − *c*_3_)+tanh(*c*_3_)] (Appendix C).

**Figure 2.**
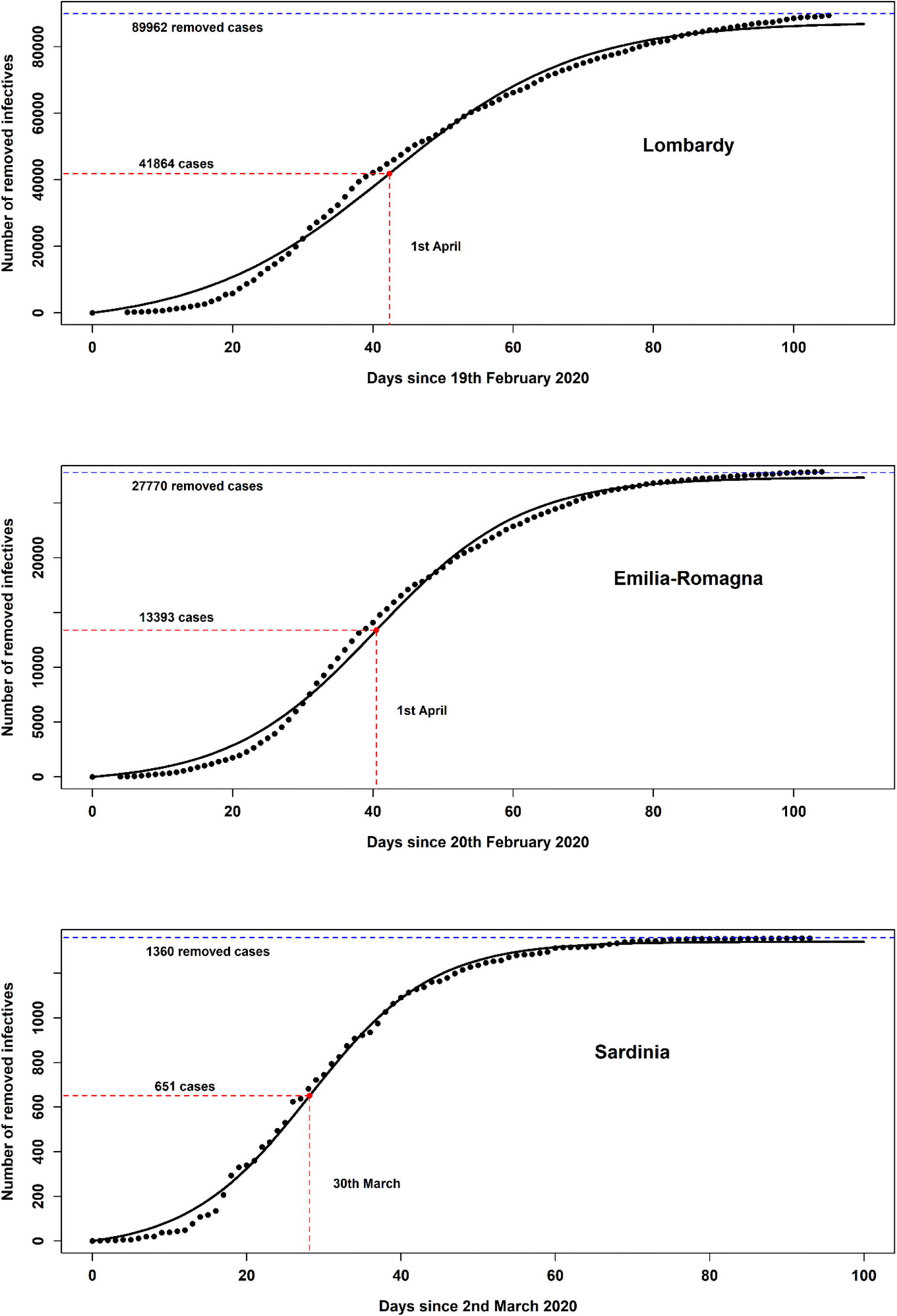
Fit of the number of removed infectives *R*(*t*) according to the Kermack-McKendrick model in three Italian regions.

Table 1 reports the main epidemic parameters of the Covid-19 epidemic in Lombardy, Emilia-Romagna and Sardinia: the basic reproduction number *R*_0_, the final numbers (for *t* → ∞) of the removed (*R*), unrecorded (*U*) and total *I_tot_* infectives, the percentages *U/I_tot_* and *R/I_tot_*, the day *t*_0_ when the epidemic started, the time *t*_peak_ (both in days, since *t*_0_, and according to calendar date) of the maximum rate *R*′(*t*_peak_) of new cases per day, with the corresponding number of removed infectives *R*(*t*_peak_), the constants *c*_1_, *c*_2_, *c*_3_ in the equation *R*(*t*) = *c*_1_·[tanh(*c*_2_*t* − *c*_3_)+tanh(*c*_3_)], determined by fitting the data on the Covid-19 epidemic with Wolfram Mathematica 12.1 [8].

**Table 1.** Main epidemic parameters from the fit of the removed infectives *R*(*t*) in Lombardy, Emilia-Romagna and Sardinia. Between brackets, we report the 95% confidence intervals.

| Parameters | Lombardy | Emilia-Romagna | Sardinia |
| --- | --- | --- | --- |
| $R_0$ | 2.07 (2.06 – 2.08) | 2.10 (2.09 – 2.11) | 2.13 (2.12 – 2.14) |
| $R(t \rightarrow \infty)$ | 87472 (84982 – 89962) | 27348 (26926 – 27770) | 1341 (1323 – 1360) |
| $U(t \rightarrow \infty)$ | 92036 (88976 – 95097) | 29619 (29022 – 30216) | 1508 (1479 – 1537) |
| $I_{tot}(t \rightarrow \infty)$ | 179508 (173957 – 185059) | 56967 (55948 – 57986) | 2849 (2802 – 2897) |
| $U/I_{tot}$ (%) | 51.3% (51.1% – 51.4%) | 52.0% (51.9% – 52.1%) | 52.9% (52.8% – 53.1%) |
| $R/I_{tot}$ (%) | 48.7% (48.6% – 48.9%) | 48.0% (47.9% – 48.1%) | 47.1% (46.9% – 47.2%) |
| $t_0$ (date) | 19<sup>th</sup> February 2020 | 20<sup>th</sup> February 2020 | 2<sup>nd</sup> March 2020 |
| $t_{peak}$ (days) | 42.3 (37.2 – 47.5) | 40.5 (36.7 – 44.2) | 28.2 (25.8 – 30.5) |
| $t_{peak}$ (date) | 1 Apr (27 Mar - 7 Apr) | 1 Apr (28 Mar - 4 Apr) | 30 Mar (28 Mar - 2 Apr) |
| $R'(t_{peak})$ | 1697 (1560 – 1835) | 669 (631 – 708) | 43 (41 – 46) |
| $R(t_{peak})$ | 41864 (40308 – 43421) | 13393 (13135 – 13650) | 651 (639 – 663) |
| $c_1$ | 4.561 (4.467 – 4.654)·10<sup>4</sup> | 1.396 (1.379 – 1.412)·10<sup>4</sup> | 6.901 (6.837 – 6.964)·10<sup>3</sup> |
| $c_2$ | 0.037 (0.035 – 0.039) | 0.048 (0.046 – 0.050) | 0.063 (0.060 – 0.066) |
| $c_3$ | 1.576 (1.478 – 1.673) | 1.942 (1.852 – 2.032) | 1.772 (1.694 – 1.850) |

Figure 3 shows the number of newly recorded infectives per day in Lombardy, Emilia-Romagna and Sardinia. These curves plot the equation 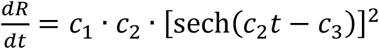 (Appendix C), which yields the rate of new removed infectives in the Kermack-McKendrick model.

**Figure 3.**
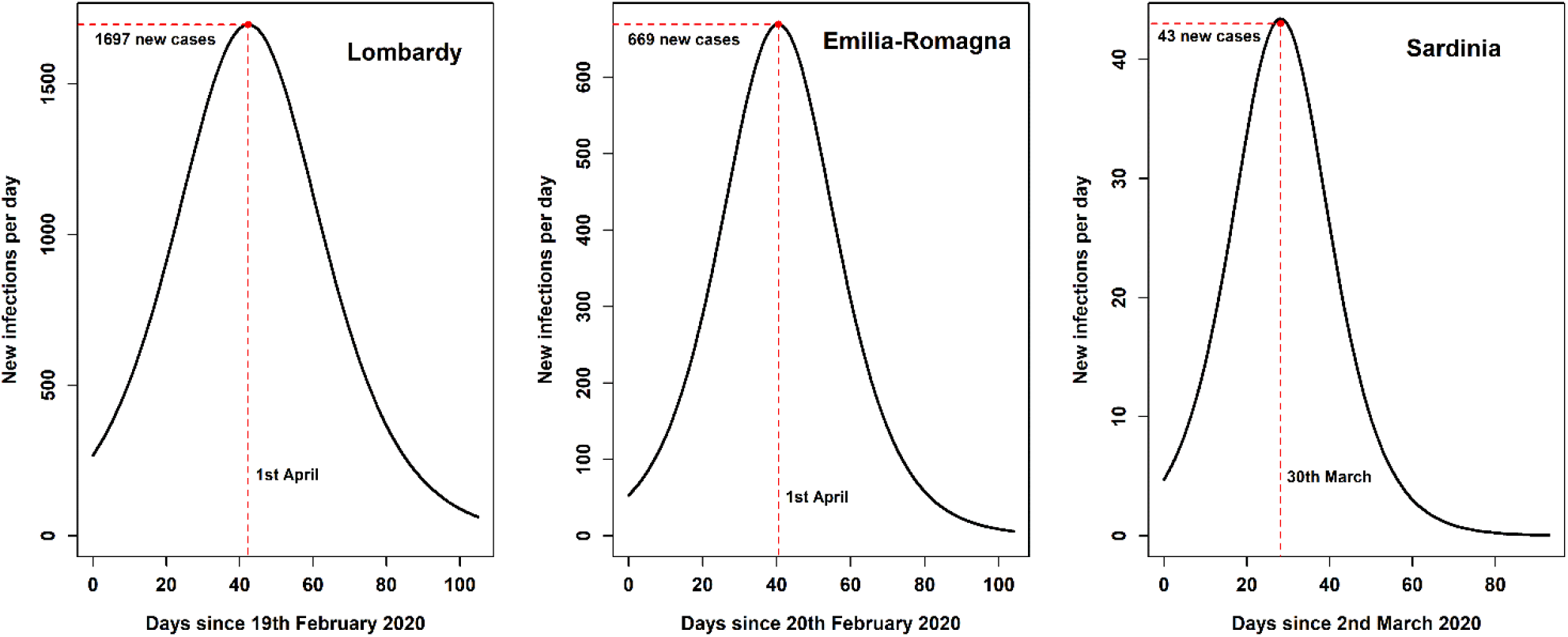
Rate 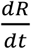 of new removed infectives per day according to the Kermack-McKendrick model in three Italian regions.

Figure 4 compares the percentages of asymptomatic infectives found in three previous investigations, conducted in Vo’ (Italy) [2], Japan [3] and China [4], with the percentage of undocumented infectives in Lombardy, Emilia-Romagna and Sardinia obtained in this study through the SIR model.

**Figure 4.**
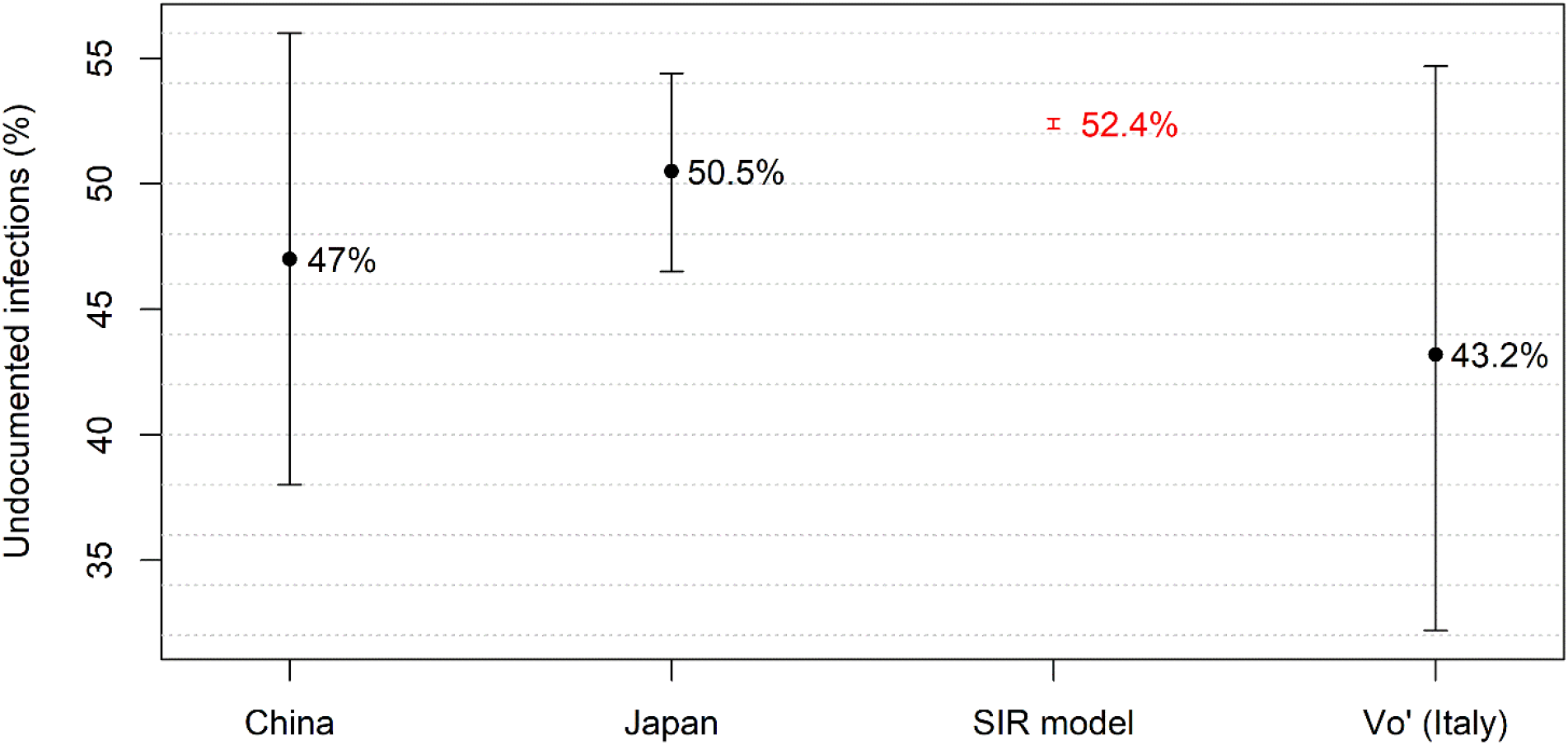
Comparison between the percentage of unrecorded infectives obtained using the SIR model and the percentages of asymptomatic infectives in three previous investigations conducted in China, Japan and Vo’ (Italy). The error bars represent the 95% confidence intervals.

The serological investigation conducted in Italy on 64660 subjects from the 15^th^ of May to the 15^th^ of July 2020 revealed that the percentage of paucisymptomatic infectives was 24.7% and that of asymptomatic infectives was 27.3%. Therefore, the total percentage of paucisymptomatic and asymptomatic infectives resulted to be 52.0%, as discussed in the preliminary report released by the Italian National Institute of Statistics [5].

The result obtained with the SIR model (shown in Figure 4) seems to be affected by a relatively small error in comparison to the errors of other studies. The reason is that the 95% confidence interval associated to our finding only represents the uncertainty intrinsic to the mathematical model, excluding the error related to the number provided by the Italian Ministry of Health [6] for removed infectives *R*(*t*) at time *t*. This number was probably understimated because of the difficulty to administer swabs or serological tests to all the suspect cases or even to subjects with overt symptoms. However, we only considered the errors associated to the statistical goodness of fit in our model, being unable to evaluate the uncertainty of the data on removed infectives.

Figure 5 shows the numbers for three Italian regions of removed (*R*), unremoved (*U*) and total (*I_tot_*) infectives, related by the equations *U* = (*R*_0_ − 1)·*R* and *I_tot_* = *R*_0_·*R*.

**Figure 5.**
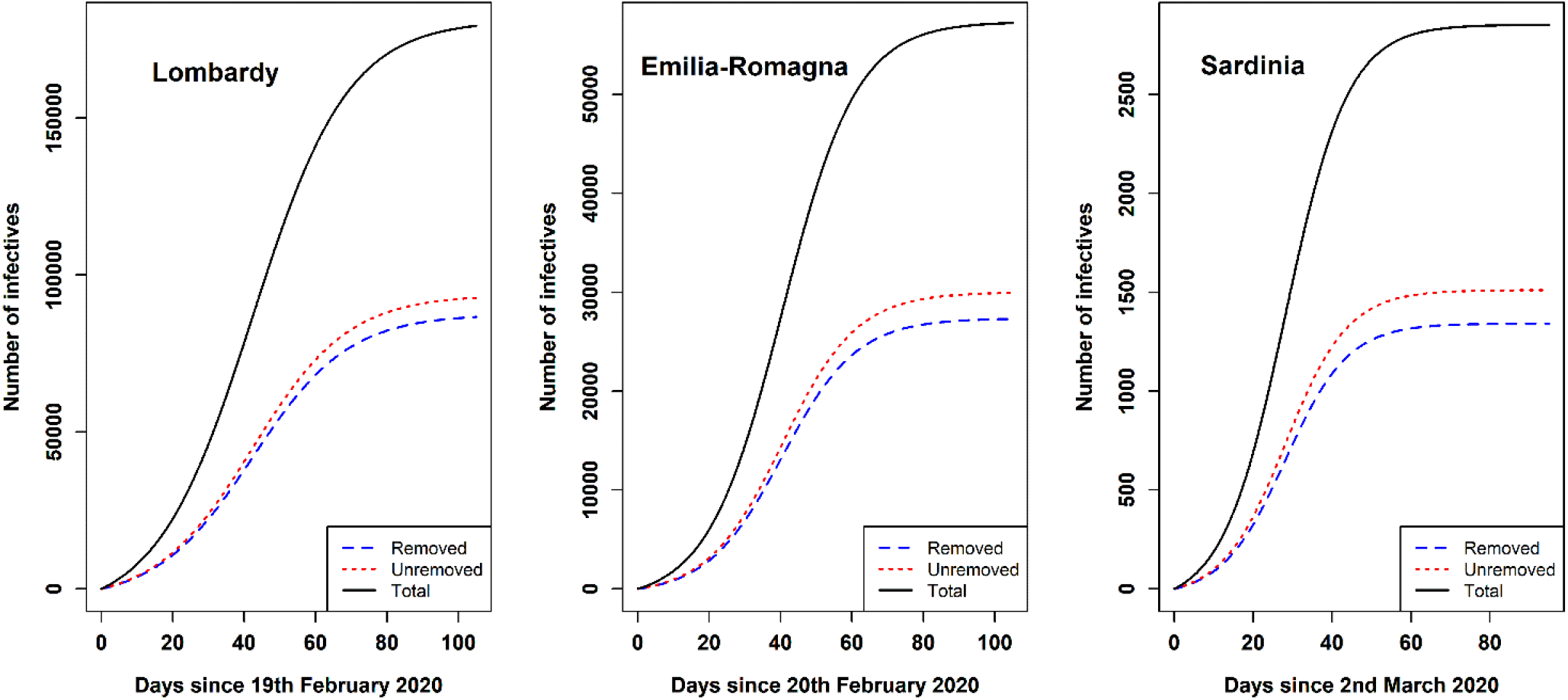
Fits of the number of removed, unrecorded and total infectives in Lombardy, Emilia-Romagna and Sardinia according to the Kermack-McKendrick model.

The assumption that the population size must be larger than the number of infected subjects corresponds to a relative error 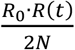 on the undocumented fraction of infectives, i.e. a percent error lower than 0.9% in Lombardy, 0.6% in Emilia-Romagna and 0.1% in Sardinia.

In Appendix D, the Kermack-McKendrick model was also used to compute the effective reproduction number *R*_eff_(*t*) and to evaluate the time corresponding to the threshold *R*_eff_(*t*) = 1 at which the epidemic starts to decline.

## Discussion

The speed at which an infection spreads is strongly influenced by the number of undocumented infected individuals who contribute to disseminate the virus without being diagnosed as positive. This study proved that in any epidemic the fraction of unrecorded infectives, compared to the total number of infections, is given by the approximated expression 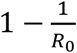, which only depends on the basic reproduction number *R*_0_.

The analytical expression of *R*_0_ found in Appendix B was exploited to compute the basic reproduction number in three Italian regions (Lombardy, Emilia-Romagna and Sardinia); the corresponding mean value *R*_0_ = 2.10 (95% confidence interval, 2.09 – 2.11) overlaps well with the result *R*_0_ = 2.2 (1.4 − 3.9) found in China [9] and the result *R*_0_ = 2.28 (2.06 – 2.52) obtained in Japan on board a cruise ship [10].

In Appendix D, the Kermack-McKendrick model was also used to compute the effective reproduction number *R*_eff_(*t*) as defined by previous authors [11].

By exploiting the aforesaid mean value of *R*_0_, we found that the percentage of unrecorded infectives was 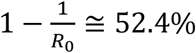 (95% confidence interval, 52.2% – 52.6%) of the total infectives.

The assumption that the population size must be larger than the number of infected subjects corresponds to a percent error lower than 1% on the undocumented fraction of infectives. As shown in Figure 4, the percentage of undocumented infectives obtained in this study overlaps well with the percentages of asymptomatic infectives found in previous investigations [2, 3, 4], confirming that the fraction of unremoved infectives is considerable and may have strong influence on the dynamics of the epidemic.

In a study conducted in Vo’ [2], a small town in Veneto (Italy), most inhabitants were tested through nasopharyngeal swabs in two consecutive surveys; the mean percentage of asymptomatic infectives corresponded to 43.2% (32.2% - 54.7%) of the total of SARS-CoV-2 infections. Important findings in this study were also that the viral load in asymptomatic infections did not significantly differ from that of symptomatic infections and that asymptomatic infectives can transmit the virus [2].

Investigation performed on the passengers of the Diamond Princess [3], a cruise ship in Yokohama (Japan), revealed that from the start of the epidemic the percentage of asymptomatic infectives on board the ship was 50.5% (46.5% - 54.4%) of the total infectives.

One of the first studies [4] to reveal the crucial role of undocumented infections in the Covid-19 pandemic estimated the undocumented fraction of infectives on the basis of a mathematical model connecting mobility data and observations of reported infections within China. The percentage of undocumented infectives turned out to be *U* = 86.2% (81.6%–89.8%) of the total number of positive cases. However, in this study the transmission rate of undocumented infectives was assumed to be *μ* = 55% (46% − 62%) of the transmission rate of symptomatic infectives [4]. On the contrary, we assumed that all infected subjects – with or without symptoms – have the same viral load, as confirmed by the investigation in Vo’ [2], and can transmit the virus at the same rate. Under this assumption, the effective percentage *U*_eff_ of undocumented infectives is given by *U*_eff_ = *μ*·*U* = 47% (38% − 56%).

Another study [12] investigated 350 attendees of a wedding in Jordan, 76 of whom tested positive for SARS-CoV-2. Among them, 36 individuals were asymptomatic, i.e. 47.4% (35.8% - 59.2%) of the total number of infected subjects.

The studies [2, 3, 12] were based on laboratory tests performed in small communities (the inhabitants of Vo’ in Italy, the passengers of a cruise ship in Japan and the attendees of a wedding in Jordan, respectively) where the Covid-19 infection had spread. On the contrary, the study in China [4] was based on a mathematical model comparing mobility data and infection diffusion in mainland China after the start of the Covid-19 epidemic.

A serological investigation in the Italian population conducted by the Italian National Institute of Statistics [5] on 64660 subjects revealed that the percentage of paucisymptomatic and asymptomatic infectives up to mid-July 2020 was 52.0%.

A Review [13] of the available evidence on asymptomatic SARS-CoV-2 infectives found that asymptomatic subjects accounted for approximately 40% to 45% of the total number of infections and could transmit the virus to others. The authors of the Review also pointed out that the high frequency of asymptomatic infections could at least partly explain the rapid spread of the virus, since infected subjects who feel and look well are likely to have more interaction with others than symptomatic infectives.

The results obtained in the aforementioned investigations [2, 3, 12, 13] concerned *asymptomatic* infected subjects, while the results found in our study included all the *undocumented* infectives, both asymptomatic subjects and paucisymptomatic or presymptomatic individuals. This can explain why the percentages of asymptomatic infected subjects in those studies [2, 3, 12, 13] turned out to be a bit lower than the percentage we found for all the undocumented infectives.

The 95% confidence intervals of the epidemiological parameters reported in Table 1 were only associated to the error intrinsic to the mathematical model considered in this study, while the uncertainty on the data concerning the removed infectives was not included, although the number of recorded positive cases was probably underestimated as a consequence of the low frequency in administering swabs and serological tests to the population in most Italian regions.

## Conclusions

Our derivation of the percentage of undocumented infectives only relied on SIR model, a cornerstone in the study of infectious disease dynamics. Despite its simplicity, SIR model describes the global dynamics of an epidemic and allows for the evaluation of several epidemiological parameters. However, more complex and realistic generalisations of the SIR model could be introduced to further refine and improve the true picture of an epidemic.

The general expression of the percentage of undocumented infectives found in this study only requires the knowledge of the basic reproduction number *R*_0_. Other methods involve numerous variables, in order to provide a more accurate description of the epidemic. However, these methods also require specific assumptions on unknown parameters of the underlying mathematical framework.

The main conclusion which can be drawn from the results obtained in this study is that unrecorded infections play a key role in the transmission of SARS-CoV-2. The high percentage of undocumented infections poses a major challenge for the control of Covid-19 and highlights the necessity to carefully monitor and adjust social distancing and other preventive measures until a vaccine is found.

## Data Availability

All the data about Covid-19 epidemic in Italy are available from the database of the
Italian Ministry of Health and Civil Protection Department

## Acknowledgments

We are grateful to Anna Maria Koopmans for translations, professional writing assistance and preparation of the manuscript.

## Appendices

### A. SIR model

The equations describing the SIR model are:

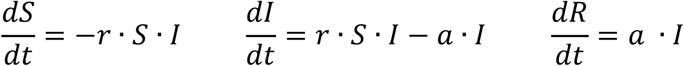

where *r* > 0 is the infection rate and *a* > 0 is the removal rate of infectives. At any time *t* the sum of *S*(*t*), *I*(*t*) and *R*(*t*) is equal to *N*, the population size:

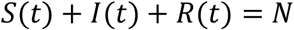

The initial conditions are:

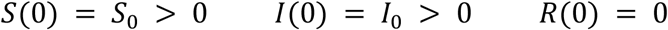

By dividing the first and third equations of the SIR model and introducing the relative removal rate *ρ* = *a/r*, one obtains

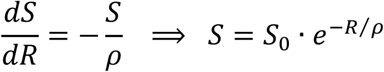

Following the Kermack-McKendrick model, if the population size is much larger than the number of infectious subjects, 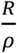 is small and *S*(*t*) can be approximated by

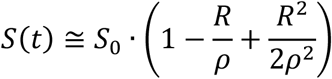

From the constraint *N* = *S*+*R*+*I* it follows that the number of infectives *I*(*t*) can be expressed as *I*(*t*) = *N* – *S*(*t*) – *R*(*t*).

The third equation of the SIR model then becomes

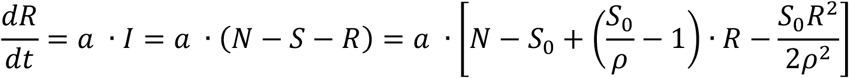

By integrating the previous equation, one obtains

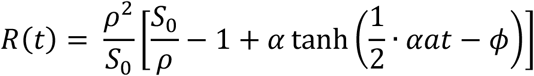

where

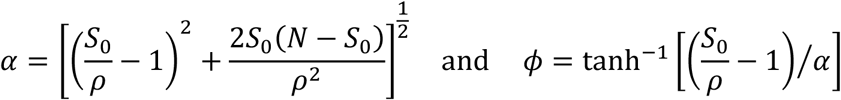

Finally, the rate 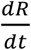 of new removed infectives per unit of time is given by

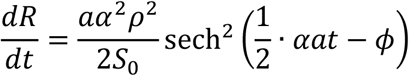

The basic reproduction number is defined as 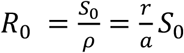. It represents, in a wholly susceptible population, the number of new infectives from one primary infection.

The SIR model assumes that the removal rate of infectives *a* and the infection rate *r* do not vary during the epidemic; consequently, the calculated curves conform roughly to the observed data. Conclusions concerning the true values of the constants *a*, *r* and *S*_0_ – as well as *R*_0_ – should not be drawn from their direct relationships with the parameters of the numerical fit.

The SIR model provides an oversimplified description of epidemic dynamics. Generalisations of it may be necessary to obtain a more accurate picture of real epidemics.

In the SIR model all the infectives *I* pass to the *R* compartment. Therefore, at any time, all the undocumented infectives are those of the *I* class: *U* = *I*. In our extended SIR model, described in the Methods section, a fraction of the infectives *I* do not pass to the removed class *R* but to an undetected class *U*_0_. It follows that the total undocumented infectives are *U* = *I* +*U*_0_. The removal rate of infectives *a* must be divided by a constant *k* > 1, corresponding to the fraction 1/*k* of the infectives *I* who actually pass to the *R* compartment: *a* → *a/k*. As a consequence, the basic reproduction number 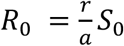 becomes *k* times greater (*R*_0_ → *kR*_0_) and the infection turns out to spread more quickly than one would expect if there were no undocumented infectives.

The equilibrium points of the system described by the differential equations of the SIR model are given, for any value of *S*, by *I* = 0. The equilibrium of the system turns out to be stable for 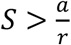 (herd immunity) and unstable for 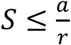. In the unstable case, a small increase in the number of infectives (*I* ≳ 0) may lead to breaking of the equilibrium and restart of the epidemic.

### B. Basic reproduction number *R*_0_

From the differential equations of the SIR model, it turns out that an epidemic occurs if *S* > *ρ*, where *ρ* is the relative removal rate and *S* is the number of susceptible subjects (with initial value *S*_0_).

The critical parameter *R*_0_ = *S*_0_/*ρ* is the basic reproduction number, representing the number of secondary infections from one primary infection in a wholly susceptible population. If *R*_0_ > 1 an epidemic ensues, if *R*_0_<1 no epidemic can occur.

From the definition of the basic reproduction number *R*_0_, it follows that each primary contagious case produces |*R*_0_ − 1| new secondary cases in a completely susceptible population. In a neighbourhood of the initial time *t* = 0, the basic reproduction number *R*_0_ can be assumed to be constant.

The number of new infectives *I*(*t*)≡*I_t_*, at any time *t* in the neighbourhood of *t* = 0, is |*R*_0_ − 1| times the number of infectives *I_t_* _− 1_ at the previous time *t* − 1, i.e. *I_t_* = |*R*_0_ − 1|·*I*_*t* − 1_.

By iterating this procedure up to the initial time *t* = 0, when *I*(0) = *I*_0_, one gets:

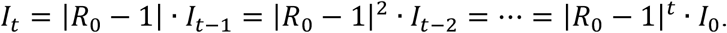

By inverting the equation *I_t_* = |*R*_0_ − 1|*^t^*·*I*_0_ and assuming *t* in a neighbourhood of *t* = 0, the basic reproduction number turns out to be:

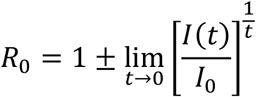

where the plus and minus signs correspond either to a growing or declining epidemic with *R*_0_ > 1 or *R*_0_<1, respectively.

Being 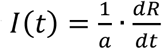 (as follows from the third equation of the SIR model, Appendix A), the previous expression of *R*_0_ can be written as

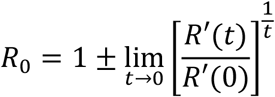

where 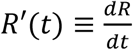 is the rate of new removed infectives per unit of time.

### C. Data fit

The number *R*(*t*) of removed infectives against time *t* can be fitted by the curve

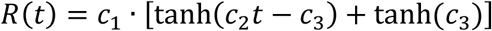

where the parameters *c*_1_, *c*_2_, *c*_3_ are related to four epidemiological characteristics: the removal rate of infectives *a*, the infection rate *r*, the initial number of susceptible subjects *S*_0_ and the population size *N*.

The initial number of removed infectives is *R*(*t* → 0) = 0, while their final number *R*(*t* → ∞) is

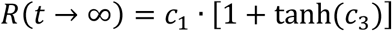

The rate of new removed infectives per unit of time is

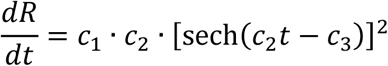

The time *t*_peak_ corresponding to the maximum of *dR/dt*, flex of the *R*(*t*) curve, is:

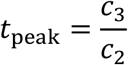

The maximum rate of new detected infections per unit of time is

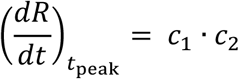

The number of removed infectives *R*(*t*) at time *t*_peak_ turns out to be

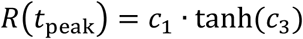

The basic reproduction number *R*_0_ is given by

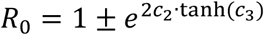

The best-fit of the Covid-19 data in Lombardy, Emilia-Romagna and Sardinia was obtained through the “NonlinearModelFit” algorithm of Wolfram Mathematica 12.1, which also provided the 95% confidence intervals of the epidemiological parameters.

The adjusted *R*-squared, measuring the goodness of fit, turned out to be about *R*_2_ = 0.999 in all the Italian regions considered in this study.

### D. Effective reproduction number

The time course of an epidemic can be described by the effective reproduction number *R*_eff_(*t*), which is defined as the average number of new secondary infected cases per primary case at time *t*.

*R*_eff_(*t*) represents the time development of the basic reproduction number *R*_0_ due to the decrease of susceptible individuals and the implementation of control measures. If *R*_eff_(*t*) < 1, the epidemic is declining and can be considered as under control; the opposite occurs if *R*_eff_(*t*) > 1.

The effective reproduction number *R*_eff_(*t*) is given by

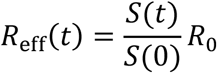

The assumption *I*_0_ ≅ 0 yields *N* ≅ *S*_0_ and *S*(*t*) ≅ *S*_0_ − *I_tot_*(*t*). The effective reproduction number can then be expressed as

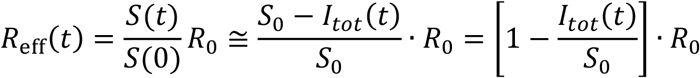

The minimum number of initial susceptible individuals *S*_0_ cannot be less than the final number of total infectives *I_tot_*(*t* → ∞) for an infection rate *r* equal to one (*r* = 1); analogously, the maximum value of *S*_0_ cannot exceed the population size *N*:

If we require that the limit of *R*_eff_(*t*) as *t* → ∞ is zero, then *S*_0_ must be assumed equal to its lower bound, *I_tot_*(*t* → ∞), and the effective reproduction number becomes

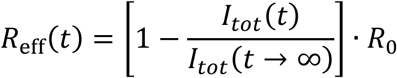

As discussed in the Methods section, *I_tot_*(*t*) ≅ *R*_0_·*R*(*t*) if the population size is much larger than the number of infected subjects; in this case the previous equation can be written as:

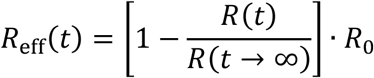

The following Figure represents the effective reproduction number *R*_eff_(*t*) against time *t* in three Italian regions: Lombardy, Emilia-Romagna and Sardinia.

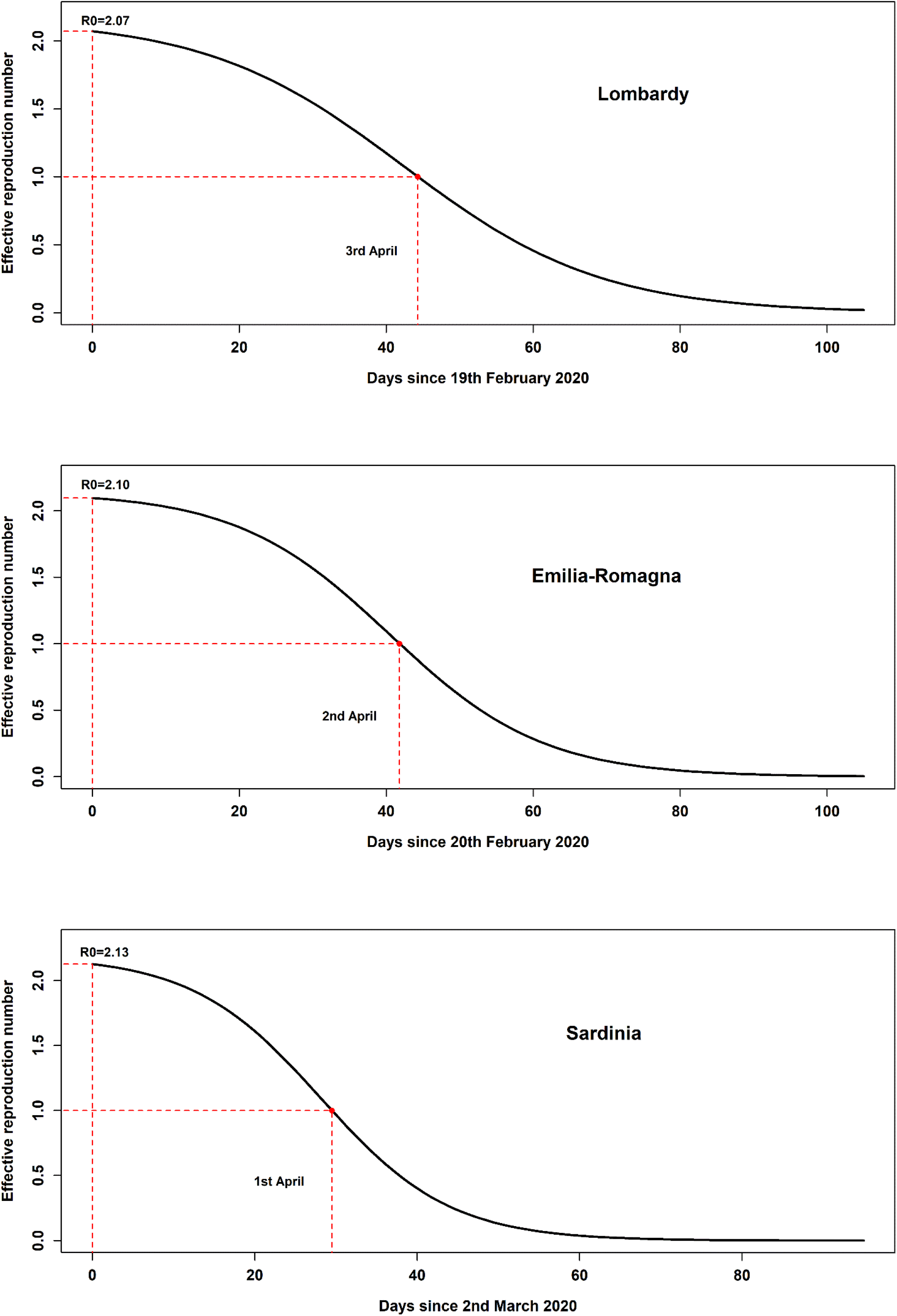

The Kermack-McKendrick model of *R*(*t*) discussed in Appendix A can be linearized in the neighbourhood of the time *t*_peak_ corresponding to the maximum of the rate 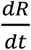 of new removed cases per unit of time: 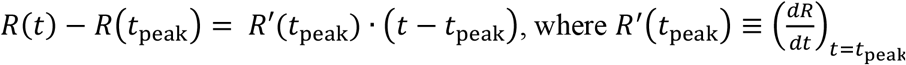.

By substituting *t* with *t*_1_, corresponding to the threshold value *R*_eff_(*t*_1_) = 1, one can compute the time difference ∆*t* = *t*_1_ − *t*_peak_:

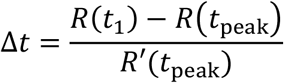

The number of removed infectives *R*(*t*_1_) at time *t* = *t*_1_ can be obtained from the equation expressing *R*_eff_(*t*) in terms of *R*(*t*):

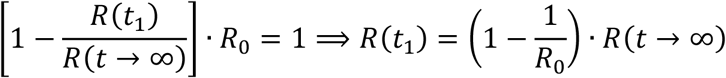

By substituting *R*(*t*_1_) into the equation of ∆*t* and expressing *R*(*t*_peak_), *R*′(*t*_peak_) and *R*(*t* → ∞) in terms of the parameters *c*_1_, *c*_2_, *c*_3_ of the *R*(*t*) fit discussed in Appendix C, the difference ∆*t* between times *t*_1_ and *t*_peak_ becomes:

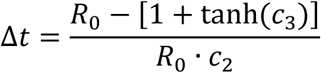

Being *t*_peak_ = *c*_3_*/c*_2_ (Appendix C), the time *t*_1_ corresponding to *R*_eff_(*t*_1_) = 1 turns out to be

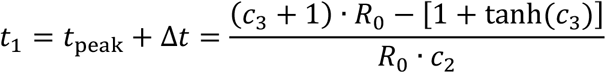

The following Table reports the 95% confidence interval for the threshold value *R*_eff_(*t*_1_) = 1 of the effective reproduction number and the corresponding time *t*_1_ (both in days, since the start of the epidemic, and according to calendar date) in the Italian regions considered in this study.

|  | **Lombardy** | **Emilia-Romagna** | **Sardinia** |
| --- | --- | --- | --- |
| $R_{\text{eff}}(t_1)$ | 1.00 (0.80 – 1.20) | 1.00 (0.81 – 1.19) | 1.00 (0.84 – 1.16) |
| $t_1$ (days) | 44.3 (39.3 – 49.4) | 41.8 (38.1 – 45.5) | 29.5 (27.2 – 31.9) |
| $t_1$ (date) | 3 Apr (29 Mar - 8 Apr) | 2 Apr (29 Mar - 6 Apr) | 1 Apr (29 Mar - 3 Apr) |

